# Impact of Recent Job Loss on Sleep, Energy Consumption and Diet

**DOI:** 10.1101/2021.10.11.21264815

**Authors:** Salma Batool-Anwar, Candace Mayer, Patricia L. Haynes, Yilin Liu, Cynthia A. Thomson, Stuart F. Quan

**Affiliations:** Division of Sleep and Circadian Disorders, Brigham and Women’s Hospital, Harvard Medical School, Boston, MA; Mel and Enid Zuckerman College of Public Health, University of Arizona, Tucson, AZ

## Abstract

To examine how sleep quality and sleep duration affect caloric intake among those experiencing involuntary job loss.

**Methods:** Adequate sleep and self reported dietary recall data from the Assessing Daily Activity Patterns through Occupational Transitions (ADAPT) study was analysed. Primary sleep indices used were total sleep time, time spent in bed after final awakening, and sleep quality as measured by the Daily Sleep Diary (DSD). Mean Energy consumption (MEC) was the primary nutritional index. Secondary indices included diet quality using the Health Eating Index 2015 (HEI), and self-reported intake of protein, carbohydrates and fats.

**Results:** The study participants were comprised mainly of women (61%) and non-Hispanic white. The participants had at least 2 years of college education and mean body mass index of 30.2±8.08 (kg/m ^2^). The average time in bed was 541.8±77.55 minutes and total sleep time was 461.1±56.49 minutes. Mean sleep efficiency was 91±6%, self-reported sleep quality was 2.40±0.57 (0-4 scale, 4 = very good), and minutes earlier than planned morning awakening were 14.36±24.15. Mean HEI score was 47.41±10.92. Although the MEC was below national average for both men and women, male sex was associated with higher MEC. In a fully adjusted model sleep quality was positively associated with MEC.

**Conclusion:** Daily overall assessments of sleep quality among recently unemployed persons were positively associated with mean energy consumption. Additionally, the diet quality of unemployed persons was found to unhealthier than the average American and consistent with the relationship between poor socioeconomic status and lower diet quality.

## Introduction

Obesity is a major public health concern; 38.3% of women and 34.3 of men ^1^ in the United States are obese. It is not only the result of low physical activity and overconsumption of high energy yielding foods, socioeconomic status also plays a major role ^2^. Obesity disproportionately affects people of lower socioeconomic status (SES) in part because their limited financial resources result in consumption of calorically dense unhealthy food. This contributes to the risk of developing obesity^3^.

A major determinant of SES is employment status. Unemployment is one indicator of a reduced SES and is associated with higher levels of stress. Unemployment includes involuntary job loss which is an important disruptive life event. It can cause additional unanticipated psychological and economic stress with the former afflicting women disproportionately^4^. Involuntary job loss is positively associated with greater symptoms of depression^5^, disruptions in daily routine changes and poor sleep quality. Unemployment has been shown to be positively associated with obesity^6,7^. In one study, women were more likely to be diagnosed with obesity after involuntary job loss^8^.

Sleep is another factor that affects obesity risk. Lower sleep quality and reductions in sleep duration have been shown to increase food intake resulting in becoming overweight or obese ^9^. Sleep deficiency can change the secretory pattern of leptin and ghrelin leading to hunger and a craving for calorically dense food^10,11^.

There are no prior studies that have investigated the impact of whether sleep quality or sleep duration influences caloric consumption in those that have experienced involuntary job loss. The Assessing Daily Activity Patterns through Occupational Transitions (ADAPT) Study is an ongoing longitudinal cohort study of individuals who have suffered involuntary job loss. We analyzed cross sectional data from the baseline assessment of the ADAPT study and hypothesized that disrupted, short sleep would be associated with increased energy intake among these individuals.

## Methods

### Participants

Study participants were part of the on-going ADAPT Study, an 18-month longitudinal study examining changes in sleep, social rhythms, and obesity following an involuntary job loss^12^. The study protocol and recruitment strategy have been described in detail previously. Briefly, all individuals who applied for unemployment insurance (UI) in the greater Tucson, Arizona and surrounding areas between October 2015 and December 2018 received study recruitment flyers within their UI intake packets. Interested individuals contacted study staff and completed phone screens assessing exclusion criteria; potentially eligible individuals were then scheduled for in-person screening visits. Individuals were eligible if they had experienced an involuntary job loss within 90 days of study enrollment, had been with their employer for at least six months, were currently employed less than 5 hours per week and did not complete any night shift work within the last 30 days. During the in-person screening, participants provided written informed consent, as well as information about their demographics, employment and medical history. They also were screened for homelessness, existing physiological and mental health conditions, substance abuse, and major sleep diagnoses which could interfere with social rhythms and sleep patterns. Those who passed screening completed validated mental health and sleep diagnostic interviews. An overnight at-home screening for sleep apnea was performed utilizing the ApneaLink Plus™ (ResMed, San Diego, CA) to exclude moderate sleep apnea as a cause of sleep disruption.

Data used in this analysis originated from the study’s baseline visit. Of the 446 adults who provided written consent, 191 participants met eligibility criteria and completed a baseline assessment visit, including an at-home data collection period lasting two weeks. Participants were considered for the current analysis if there was an acceptable assessment of sleep and diet on their sleep diaries and dietary recalls respectively for analysis. However, 8 participants were excluded as outliers because their mean energy consumption (MEC) was significantly less than commonly reported norms ^13,14^. Descriptive statistics for the study sample are reported in Table 1.

**Table 1:** Baseline Characteristics

| VARIABLES | N | Mean | sd |
| --- | --- | --- | --- |
| *Demographic/Anthropometric* |  |  |  |
| Age (*years*) | 183 | 40.93 | 10.24 |
| Gender (% *Men*) | 183 | 38.8 | -- |
| Ethnicity (% *Hispanic*) | 183 | 33.0 | -- |
| Education | 183 | 4.027 | 1.61 |
| BMI (kg/m<sup>2</sup>) | 183 | 30.22 | 8.08 |
| *Sleep* |  |  |  |
| Time in Bed (*min*) | 183 | 541.80 | 77.55 |
| Total Sleep Time (*min*) | 183 | 461.10 | 56.49 |
| Sleep Latency (*min*) | 183 | 26.56 | 22.52 |
| Sleep Efficiency (%) | 183 | 0.91 | 0.06 |
| Wake After Sleep Onset Episodes | 183 | 1.52 | 1.09 |
| Early morning awakening (*min*) | 183 | 14.36 | 24.15 |
| Sleep quality | 183 | 2.40 | 0.57 |

### Measures

#### Demographic and Anthropometric

Age, ethnicity and biological sex were collected during the initial interview. Height and weight were measured to calculate the body mass index (BMI kg/m^2^).

#### Diet Assessment

During the two-week, at-home baseline data collection period, participants completed up to three 24-hour dietary recalls administered by trained diet assessors at the Behavioral Measurements and Interventions Shared Resource of the University of Arizona Cancer Center utilizing the gold-standard USDA Multi-pass Dietary recall ^15^ and the Nutrient Database System of the University of Minnesota for nutrient analysis^16^ These interviews were supported by the Remote Food Photography Method^17^, in which participants took pictures of all food and beverages prior to consumption, as well as after they had finished eating and drinking. Photos were used to review recall as a final verification of the multi-pass data. The diet recalls provided information on the types and quantity of food, including energy and nutrient values. At least 3 dietary recalls were completed by 172 participants (95.6% of the cohort). The primary index for this analysis was MEC (kcal). Secondary indices included diet quality estimated using the Health Eating Index 2015 (HEI)^18^ using standardized approaches to score, and self-reported intake of protein, carbohydrates and fats.

#### Sleep Measures

Sleep variables of interest were measured using the valid and reliable Daily Sleep Diary (DSD)^19^, the recommended subjective sleep assessment instrument of the insomnia research consensus panel. Upon wakening from their sleep, participants completed the DSD via mobile application. The DSD was completed at least 15 times by 170 participants with only 3 participants completing less than 4 days of diary data. The primary indices of interest were total sleep time (TST) and minutes earlier than desired morning awakening (EMA) as indices of sleep duration. Additionally, sleep onset latency, sleep efficiency, number of wake after sleep onset episodes and time in bed were calculated. Self-reported sleep quality was assessed using the 5 point Likert scale incorporated into the DSD ^19^.

### Statistical Analysis

For baseline characteristics, mean (SD) for continuous variables and percentages for categorical variables were calculated. After removing the extreme outliers, we fitted linear regression models to predict MEC and HEI with sleep indices as predictors. Finally, five models were performed in multiple regression analysis. These models were adjusted for age, gender, education level, and body mass index. The results from regression analysis are presented as β-coefficients (standardized/unstandardized) with *p* values. In Model 1 we included energy consumption as predicted by total sleep time. In Model 2 we included energy consumption as predicted by time in bed. In Model 3, we included energy consumption as predicted by early morning awakenings. In Model 4, we included energy consumption as predicted by sleep quality. In our final Model 5, we included energy consumption as predicted by all sleep indicies combined from other models. The level of statistical significance for all models was set at 0.05. To test the robustness of the analysis, we conducted a sensitivity analysis by excluding 3 participants who did not completed the DSD at least four times. All statistical analyses were done using STATA version 11 (StataCorp, LLC, College Station, TX, USA).

## Results

The demographic and anthropometric characteristics of the study population are described in Table 1. Most participants were women (61%) and non-Hispanic white with the remainder primarily Hispanic. On average, study participants had at least 2 years of college education and had a mean body mass index of 30.22 (kg/m^2^)

As shown on Table 1, average time in bed (TIB) and total sleep time (TST) were 541.8 minutes (9.0 hrs) and 461.1 minutes (7.6 hrs) respectively. However, the range was large with minimums of [5.7 hrs for TIB/ 5.2 hrs for TST] and maximums of [ 15.2 hrs for TIB and 10.1 for TST] respectively. Mean sleep efficiency was within normal limits (91%). There generally were few episodes of wake after sleep onset, and only a small amount of time (14 min) spent awake in the morning (EMA). Self-reported sleep quality was 2.39 using a 5-point Likert scale from 0 to 4 representing very poor, poor, good and very good sleep respectively.

Table 2 shows the MEC as well as the intake of protein, carbohydrates, fats, fruits, and vegetables. Normative values for men and women in the United States are provided for comparison. The MEC was below averages for national data in both men and women^20-22^. Absolute intake of protein and carbohydrates was higher than recommended levels but was within recommended levels as a percent of energy consumption. Absolute intake of total fat was at the higher limit of recommended levels but exceeded them as a percent of energy consumption. Consumption of fruit and vegetables were below recommendations. The HEI also was markedly reduced (Men: 43.9±8.9; Women: 49.9±10.9 vs. 59 for average American diet)^21^.

**Table 2:** Self-reported 24-hour Dietary Intake among Recently Unemployed U.S. Adults

| VARIABLES | Men<br>(N=70) | sd | Women<br>(N=110) | sd | Recommended or Average Values |
| --- | --- | --- | --- | --- | --- |
| Mean energy consumption (kcal) | 1,898.5<sup>7</sup> | 654.7 | 1522.2<sup>7</sup> | 501.2 | 2000-2400 (Men); 1600-2000(Women)<sup>1</sup> |
| Carbohydrates (g) | 220.7<sup>7</sup> | 94.1 | 175.9<sup>7</sup> | 70.2 | 130 (Men and Women)<sup>1</sup> |
| *Carbohydrates (% of Energy)*<sup>8</sup> | 44.9 | 11.6 | 45.1 | 9.4 | 45-65 (Men and Women)<sup>1</sup> |
| Protein (g) | 78.8<sup>7</sup> | 33.5 | 60.4<sup>7</sup> | 19.5 | 56 (Men); 46 (Women)<sup>1</sup> |
| *Protein (% of Energy)*<sup>8</sup> | 16.8 | 5.3 | 16.4 | 4.6 | 10-35 (Men and Women)<sup>1</sup> |
| Total Fat (g)<sup>8</sup> | 77.7 | 30.6 | 65.0 | 26.8 | 44-77<sup>5</sup> |
| *Total Fat (% of Energy)*<sup>8</sup> | 35.4 | 8.3 | 36.9 | 7.7 | 20-35 (Men and Women)<sup>1</sup> |
| Saturated Fatty Acids (g)<sup>8</sup> | 26.7 | 12.9 | 22.4 | 11.4 | As low as possible<sup>3</sup> |
| Trans-fatty Acids (g)<sup>8</sup> | 2.1 | 1.3 | 1.6 | 1.0 | As low as possible<sup>3</sup> |
| Daily servings of Fruits (cups eq/d) | 1.3<sup>7</sup> | 1.4<sup>7</sup> | 0.8 | 0.7 | 2.5 (Men) 1.5 (Women)<sup>2</sup> |
| Daily servings of Vegetables (cups eq/d) | 2.6<sup>7</sup> | 1.4<sup>7</sup> | 3.2 | 1.3 | 3.5 (Men); 2.5 (Women)<sup>2</sup> |
| Healthy Eating Index | 43.9<sup>7</sup> | 8.9 | 49.9<sup>7</sup> | 10.9 | Highest Score: 100; Average American Diet: 59<sup>6</sup> |

<sup>1</sup> U.S. Department of Agriculture and U.S. Department of Health and Human Services. Dietary Guidelines for Americans, 2020-2025. 9th Edition. December 2020. Available at DietaryGuidelines.gov. Based on age ≥19 years
<sup>2</sup> *ibid*, Based on age 31-59 years
<sup>3</sup> Institute of Medicine 2005. Dietary Reference Intakes for Energy, Carbohydrate, Fiber, Fat, Fatty Acids, Cholesterol, Protein, and Amino Acids. Washington, DC: The National Academies Press. https://doi.org/10.17226/10490.
<sup>4</sup> 2015-2020 Dietary Guidelines for Americans. U.S. Department of Health and Human Services and U.S. Department of Agriculture. https://health.gov/dietaryguidelines/2015/guidelines/.
<sup>5</sup> Extrapolated from 2000 kcal diet with 20-35% fat content
<sup>6</sup> U.S. Department of Agriculture and U.S. Department of Health and Human Services. Dietary Guidelines for Americans, 2020-2025. 9th Edition. December 2020. Available at DietaryGuidelines.gov. Based on Age 31-59 years
<sup>7</sup> $p < 0.001$ vs. Recommended or Average Values
<sup>8</sup> Statistical comparison not performed because Recommended Values were a range

Univariate regression models examining the impact of age, gender, education, and BMI on MEC are presented in Table 3. Male gender was associated with higher MEC but age, education and BMI were not. Univariate regression models assessing the effect of the various sleep variables demonstrated that only TST, TIB, EMA and sleep quality significantly affected MEC (data not shown). Therefore, each of these factors were included by themselves in multivariate regression models that also incorporated age, gender, education and BMI (Table 3). Sleep quality was positively associated with MEC while EMA was negatively associated. There was no significant relationship between MEC and TST. A final model integrating TST, TIB, EMA and sleep quality showed that only sleep quality was associated with higher MEC, but EMA had no significant impact. In a sensitivity analysis, we excluded the participants who had not completed the DSD at least 4 times. However, this did not materially change the results.

**Table 3:**
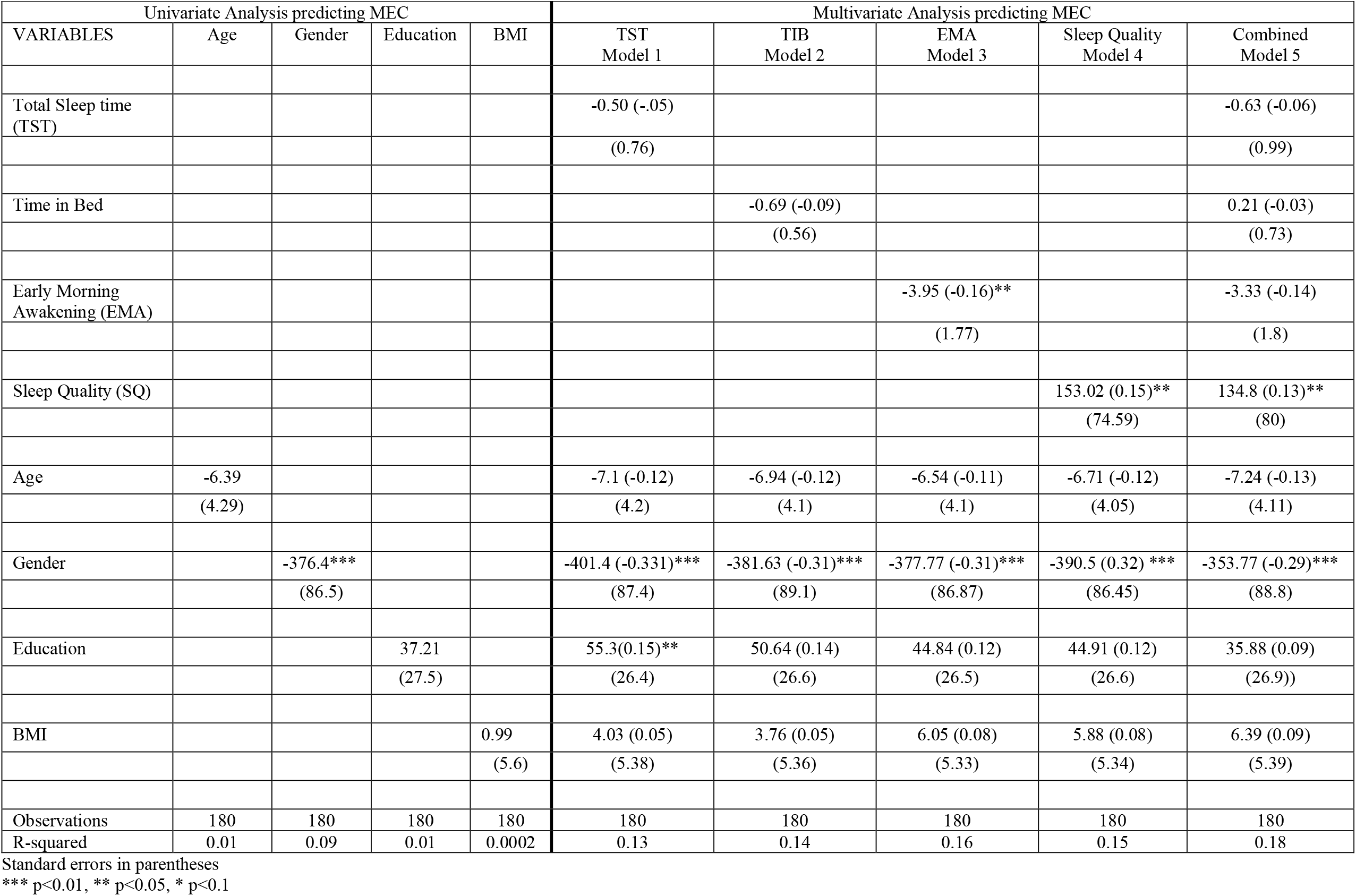
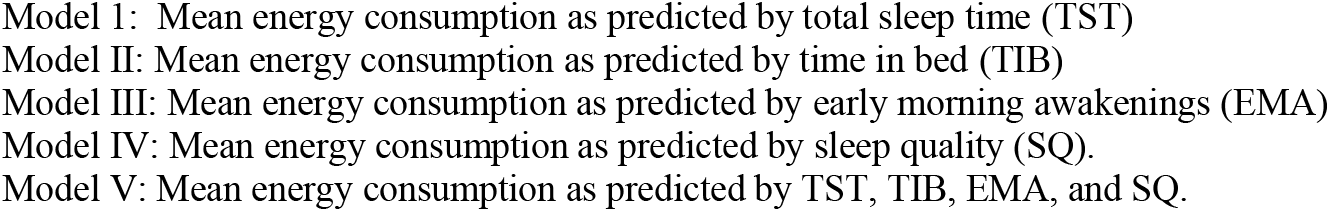
Regression Coefficients Predicting Energy Consumption

Regression models were calculated to examine the impact of sleep on dietary components and the HEI. None were shown to be significant (data not shown).

## Discussion

In this study of persons who recently involuntarily became unemployed, we did not find any significant associations between their MEC and various parameters related to sleep duration and sleep fragmentation. However, overall positive subjective sleep quality was associated with *greater* MEC. Individual dietary components and the HEI also were not related to sleep duration or fragmentation but did indicate that the diet of involuntarily unemployed persons is of lower quality, based on HEI 2015 scoring, than average for US adults.^18 21^

In most, but not all studies, sleep duration has been shown to be inversely associated with MEC. In contradistinction, our analyses did not find any significant relationship with respect to either TST or TIB. Although minutes spent awake in the morning before getting out of bed (EMA) was inversely associated with MEC, this association was borderline and not significant in a fully adjusted model. Similarly, sleep efficiency and number of wake after sleep onset episodes were not related to MEC. Explanations for increased MEC with restricted sleep duration or fragmentation include but are not limited to changes in the relative levels of satiety and hunger hormones, greater available time to eat, altered timing of meals and hedonic feeding^23^ Our data suggest that in this population, the impact of these factors is not sufficient enough to alter MEC. Importantly, a large body of evidence suggests under-reporting of dietary intake is associated with obesity, female sex and lower education^24^ and may be more common among Hispanics^25^ who accounted for 33% of our sample. Systematic under-reporting of intake may have undermined our ability to capture significant associations between energy intake and sleep in this study.

Subjective sleep quality was positively correlated with MEC; better sleep quality was associated with higher levels of MEC. The direction of this finding is inconsistent with previous studies that have noted better sleep quality is associated with more nutritious diets and less obesity^26 27^. The lack of agreement between daily subjective overall sleep quality, and specific individual subjective sleep quality metrics as well as objective sleep quality instruments (e.g., actigraphy) has been reported previously. In a recent study of sleep quality in older adults, the specific measures assessed by the DSD used in this study was compared to the Pittsburgh Sleep Quality Index as well as subjective sleep quality recorded in the diary. Little agreement was observed among all three measures.^28^ Furthermore, subjective estimates of sleep or alertness have been shown to be a poor predictor of other aspects of human behavior and performance.^29,30^ Our findings provide a unique perspective on the use of the DSD, an instrument that is considered a gold-standard for the assessment of sleep in persons with insomnia subject to less retrospective recall bias that global estimates of sleep quality.

We observed that the diet of recently unemployed persons differed in many categories from recommendations and guidelines made by the Institute of Medicine and the US Department of Agriculture. Additionally, the HEI of the average American is already suboptimal at 59 and mean scores appear to be even lower in our sample of unemployed individuals. Food cost is inversely correlated with diet quality and is one factor that contributes to the higher prevalence of unhealthy diets in those with lower socioeconomic status.^31^ Our findings extend these previous observations by demonstrating the adverse economic impact of recent job loss is associated with worse diet quality.

In conclusion, in recently unemployed persons, subjective diary assessments of sleep quality were not associated with mean energy consumption. However, the diet quality of unemployed persons was found to unhealthier than the average American and consistent with the relationship between poor socioeconomic status and lower diet quality.

## Data Availability

Data availability upon request

## Acknowledgements

The authors would like to thank the staff and participants of the Assessing Daily Activity Patterns Through Occupational Transitions Study (ADAPT). The authors would like to gratefully acknowledge the assistance of the Arizona Department of Economic Security in study recruitment, and the support of the University of Arizona Collaboratory for Metabolic Disease Prevention and Treatment.

The ADAPT study was supported by the US National Heart, Lung, and Blood Institute (HL117995).

## Abbreviations

SES: Socioeconomic status
ADAPT: Assessing Daily Activity Patterns through Occupational Transitions
UI: unemployment insurance
MEC: mean energy consumption
BMI: body mass index
USDA: United States Department of Agriculture
HEI: Healthy Eating Index
DSD: Daily Sleep Diary
TST: total sleep time
EMA: earlier than desired morning awakening
SD: Standard Deviation
TIB: time in bed

